# Pathway-Specific Polygenic Scores for Lithium Response for Predicting Clinical Lithium Treatment Response in Patients with Bipolar Disorder

**DOI:** 10.1101/2025.03.20.25324216

**Authors:** Nigussie T. Sharew, Scott R. Clark, Sergi Papiol, Urs Heilbronner, Franziska Degenhardt, Janice M. Fullerton, Liping Hou, Tatyana Shekhtman, Mazda Adli, Nirmala Akula, Kazufumi Akiyama, Raffaella Ardau, Bárbara Arias, Roland Hasler, Hélène Richard-Lepouriel, Nader Perroud, Lena Backlund, Abesh Kumar Bhattacharjee, Frank Bellivier, Antonio Benabarre, Susanne Bengesser, Joanna M. Biernacka, Armin Birner, Cynthia Marie-Claire, Pablo Cervantes, Hsi-Chung Chen, Caterina Chillotti, Sven Cichon, Cristiana Cruceanu, Piotr M. Czerski, Nina Dalkner, Maria Del Zompo, J. Raymond DePaulo, Bruno Étain, Stephane Jamain, Peter Falkai, Andreas J. Forstner, Louise Frisen, Mark A. Frye, Sébastien Gard, Julie S. Garnham, Fernando S. Goes, Maria Grigoroiu-Serbanescu, Andreas J. Fallgatter, Sophia Stegmaier, Thomas Ethofer, Silvia Biere, Kristiyana Petrova, Ceylan Schuster, Kristina Adorjan, Monika Budde, Maria Heilbronner, Janos L. Kalman, Mojtaba Oraki Kohshour, Daniela Reich-Erkelenz, Sabrina K. Schaupp, Eva C. Schulte, Fanny Senner, Thomas Vogl, Ion-George Anghelescu, Volker Arolt, Udo Dannlowski, Detlef E. Dietrich, Christian Figge, Markus Jäger, Fabian U. Lang, Georg Juckel, Carsten Konrad, Jens Reimer, Max Schmauß, Andrea Schmitt, Carsten Spitzer, Martin von Hagen, Jens Wiltfang, Jörg Zimmermann, Till F.M. Andlauer, Andre Fischer, Felix Bermpohl, Philipp Ritter, Silke Matura, Anna Gryaznova, Irina Falkenberg, Cüneyt Yildiz, Tilo Kircher, Julia Schmidt, Marius Koch, Kathrin Gade, Sarah Trost, Ida S. Haussleiter, Martin Lambert, Anja C. Rohenkohl, Vivien Kraft, Paul Grof, Ryota Hashimoto, Joanna Hauser, Stefan Herms, Per Hoffmann, Esther Jiménez, Jean-Pierre Kahn, Layla Kassem, Po-Hsiu Kuo, Tadafumi Kato, John Kelsoe, Sarah Kittel-Schneider, Ewa Ferensztajn-Rochowiak, Barbara König, Ichiro Kusumi, Gonzalo Laje, Mikael Landén, Catharina Lavebratt, Marion Leboyer, Susan G. Leckband, Alfonso Tortorella, Mirko Manchia, Lina Martinsson, Michael J. McCarthy, Susan McElroy, Francesc Colom, Vincent Millischer, Marina Mitjans, Francis M. Mondimore, Palmiero Monteleone, Caroline M. Nievergelt, Markus M. Nöthen, Tomas Novák, Claire O’Donovan, Norio Ozaki, Andrea Pfennig, Claudia Pisanu, James B. Potash, Andreas Reif, Eva Reininghaus, Guy A. Rouleau, Janusz K. Rybakowski, Martin Schalling, Peter R. Schofield, Barbara W. Schweizer, Giovanni Severino, Paul D. Shilling, Katzutaka Shimoda, Christian Simhandl, Claire M. Slaney, Alessio Squassina, Thomas Stamm, Pavla Stopkova, Mario Maj, Gustavo Turecki, Eduard Vieta, Julia Veeh, Biju Viswanath, Stephanie H. Witt, Adam Wright, Peter P. Zandi, Philip B. Mitchell, Michael Bauer, Martin Alda, Marcella Rietschel, Francis J. McMahon, Thomas G. Schulze, Bernhard T. Baune, Klaus Oliver Schubert, Azmeraw T. Amare

**Affiliations:** Discipline of Psychiatry, School of Medicine, University of Adelaide, Adelaide, SA, Australia; Asrat Woldeyes Health Science Campus, Debre Berhan University, Debre Berhan, Ethiopia; Institute of Psychiatric Phenomics and Genomics (IPPG), University Hospital, LMU Munich, Munich, Germany; Department of Psychiatry and Psychotherapy, University Hospital, Ludwig-Maximilian-University Munich, Munich, Germany; Institute of Human Genetics, University of Bonn, School of Medicine & University Hospital Bonn, Bonn, Germany; Department of Child and Adolescent Psychiatry, Psychosomatics and Psychotherapy, LVR Klinikum Essen, University of Duisburg-Essen, Rheinische Kliniken, Essen, Germany; Neuroscience Research Australia, Sydney, NSW, Australia; School of Biomedical Sciences, University of New South Wales, Sydney, NSW, Australia; Intramural Research Program, National Institute of Mental Health, National Institutes of Health, US Department of Health & Human Services, Bethesda, MD, USA; Department of Psychiatry, University of California San Diego, San Diego, CA, USA; Department of Psychiatry and Psychotherapy, Charité - Universitätsmedizin Berlin, Campus Charité Mitte, Berlin, Germany; Department of Biological Psychiatry and Neuroscience, Dokkyo Medical University School of Medicine, Mibu, Tochigi, Japan; Unit of Clinical Pharmacology, Hospital University Agency of Cagliari, Cagliari, Italy; Unitat de Zoologia i Antropologia Biològica (Dpt. Biologia Evolutiva, Ecologia i Ciències Ambientals), Facultat de Biologia and Institut de Biomedicina (IBUB), University of Barcelona, CIBERSAM, Barcelona, Spain; Department of Psychiatry, Mood Disorders Unit, HUG - Geneva University Hospitals, Geneva, Switzerland; Department of Molecular Medicine and Surgery, Karolinska Institute, Stockholm, Sweden; Center for Molecular Medicine, Karolinska University Hospital, Stockholm, Sweden; Department of Psychiatry, University of California San Diego, San Diego, CA, United States; INSERM UMR-S 1144, Université Paris Cité, Département de Psychiatrie et de Médecine Addictologique, AP-HP, Groupe Hospitalier Saint-Louis-Lariboisière-F.Widal, Paris, France; Bipolar and Depressive Disorders Program, Institute of Neuroscience, Hospital Clinic, University of Barcelona, IDIBAPS, CIBERSAM, Barcelona, Catalonia, Spain; Department of Psychiatry and Psychotherapeutic Medicine, Research Unit for bipolar affective disorder, Medical University of Graz, Graz, Austria; Department of Quantitative Health Sciences, Mayo Clinic, Rochester, MN, United States; Department of Psychiatry and Psychology, Mayo Clinic, Rochester, MN, United States; Université Paris Cité, Inserm, Optimisation Thérapeutique en Neuropsychopharmacologie, F-75006, Paris, France; The Neuromodulation Unit, McGill University Health Centre, Montreal, Canada; Department of Psychiatry & Center of Sleep Disorders, National Taiwan University Hospital, Taipei, Taiwan; Human Genomics Research Group, Department of Biomedicine, University Hospital Basel, Switzerland; Institute of Medical Genetics and Pathology, University Hospital Basel, Basel, Switzerland; Institute of Neuroscience and Medicine (INM-1), Research Center Jülich, Jülich, Germany; Douglas Mental Health University Institute, McGill University, Montreal, Canada; Psychiatric Genetic Unit, Poznan University of Medical Sciences, Poznan, Poland; Department of Biomedical Sciences, University of Cagliari, Cagliari, Italy; Department of Psychiatry and Behavioral Sciences, Johns Hopkins University, Baltimore, MD, United States; Inserm U955, Translational Psychiatry laboratory, Fondation FondaMental, Créteil, France; Max Planck Institute of Psychiatry, Munich, Germany; Centre for Psychiatry Research, Department of Clinical Neuroscience, Karolinska Institute, Stockholm, Sweden; Pôle de Psychiatrie Générale Universitaire, Hôpital Charles Perrens, Bordeaux, France; Department of Psychiatry, Dalhousie University, Halifax, Nova Scotia, Canada; Biometric Psychiatric Genetics Research Unit, Alexandru Obregia Clinical Psychiatric Hospital, Bucharest, Romania; University Department of Psychiatry and Psychotherapy Tuebingen, University of Tübingen, Tuebingen, Germany; Department of General Psychiatry, University of Tuebingen, Tuebingen, Germany; Department of Biomedical Resonance, University of Tuebingen, Tuebingen, Germany; Department of Psychiatry, Psychotherapy and Psychosomatics, University Hospital of Frankfurt, Goethe University, Frankfurt, Germany; Department of Psychiatry and Psychotherapie, University of Bern, Switzerland; Department of Immunology, Faculty of Medicine, Ahvaz Jundishapur University of Medical Sciences, Ahvaz, Iran; Department of Psychiatry and Psychotherapy, Mental Health Institute Berlin, Berlin, Germany; Institute for Translational Psychiatry, University of Münster, Münster, Germany; AMEOS Clinical Center Hildesheim, Hildesheim, Germany; Center for Systems Neuroscience (ZSN), Hannover, Germany; Karl-Jaspers Clinic, European Medical School Oldenburg-Groningen, Oldenburg, 26160, Germany; Department of Psychiatry II, Ulm University, Bezirkskrankenhaus Günzburg, Günzburg, Germany; Department of Psychiatry, Ruhr University Bochum, LWL University Hospital, Bochum, Germany; Department of Psychiatry and Psychotherapy, Agaplesion Diakonieklinikum, Rotenburg, Germany; Department of Psychiatry and Psychotherapy, University Medical Center Hamburg-Eppendorf, Hamburg, Germany; Department of Psychiatry, Health North Hospital Group, Bremen, Germany; Department of Psychiatry and Psychotherapy, Bezirkskrankenhaus Augsburg, Augsburg, Germany; Laboratory of Neuroscience (LIM27), Institute of Psychiatry, University of Sao Paulo, São Paulo, Brazil; Department of Psychosomatic Medicine and Psychotherapy, University Medical Center Rostock, Rostock, Germany; Clinic for Psychiatry and Psychotherapy, Clinical Center Werra-Meißner, Eschwege, Germany; Department of Psychiatry and Psychotherapy, University Medical Center Göttingen, Göttingen, Germany; German Center for Neurodegenerative Diseases (DZNE), University of Göttingen, Göttingen, Germany; Psychiatrieverbund Oldenburger Land gGmbH, Karl-Jaspers-Klinik, Bad Zwischenahn, Germany; Department of Neurology, University Hospital rechts der Isar, School of Medicine, Technical University of Munich, Munich, Germany; Department of Psychiatry and Psychotherapy, University Hospital Carl Gustav Carus, Medical Faculty, Technische Universität Dresden, Germany; Department of Psychiatry and Psychotherapy, Philipps-University, Marburg, Germany; Institute for Medical Informatics, University Medical Center Göttingen, Göttingen, Germany; Mood Disorders Center of Ottawa, Ontario, Canada; Department of Pathology of Mental Diseases, National Institute of Mental Health, National Center of Neurology and Psychiatry, 4-1-1 Ogawahigashi, Kodaira, Tokyo 187-8553, Japan; Service de Psychiatrie et Psychologie Clinique, Centre Psychothérapique de Nancy - Université de Lorraine, Nancy, France; Department of Public Health & Institute of Epidemiology and Preventive Medicine, College of Public Health, National Taiwan University, Taipei, Taiwan; Laboratory for Molecular Dynamics of Mental Disorders, RIKEN Brain Science Institute, Saitama, Japan; Department of Psychiatry, Psychosomatic Medicine and Psychotherapy, University Hospital Frankfurt, Frankfurt, Germany; Department of Psychiatry, Psychotherapy and Psychosomatic Medicine, University Hospital of Würzburg, Wurzburg, Germany; Department of Adult Psychiatry, Poznan University of Medical Sciences, Poznan, Poland; Department of Psychiatry and Psychotherapeutic Medicine, Landesklinikum Neunkirchen, Neunkirchen, Austria; Department of Psychiatry, Hokkaido University Graduate School of Medicine, Sapporo, Japan; Institute of Neuroscience and Physiology, the Sahlgrenska Academy at the Gothenburg University, Gothenburg, Sweden; Department of Medical Epidemiology and Biostatistics, Karolinska Institutet, Stockholm, Sweden; Inserm U955, Translational Psychiatry laboratory, Université Paris-Est-Créteil, Department of Psychiatry and Addictology of Mondor University Hospital, AP-HP, Fondation FondaMental, Créteil, France; Office of Mental Health, VA San Diego Healthcare System, San Diego, CA, United States; Department of Psychiatry, University of Perugia, Italy; Section of Psychiatry, Department of Medical Sciences and Public Health, University of Cagliari, Cagliari, Italy; Department of Pharmacology, Dalhousie University, Halifax, NS, Canada; Department of Clinical Neurosciences, Karolinska Institute, Stockholm, Sweden; Department of Psychiatry, VA San Diego Healthcare System, San Diego, CA, United States; Department of Psychiatry, Lindner Center of Hope / University of Cincinnati, Mason, OH, United States; Mental Health Research Group, IMIM-Hospital del Mar, Barcelona, Catalonia, Spain; Department of Genetics, Microbiology and Statistics, Faculty of Biology, University of Barcelona, Barcelona, Spain; Department of Psychiatry and Psychotherapy, Medical University of Vienna, Vienna, Austria; Centro de Investigación Biomédica en Salud Mental (CIBERSAM), Instituto de Salud Carlos III, Madrid, Spain; Institut de Biomedicina de la Universitat de Barcelona (IBUB), Barcelona, Spain; Institut de Recerca Sant Joan de Déu (IRSJD), Esplugues de Llobregat, Spain; Neurosciences Section, Department of Medicine, Surgery and Dentistry “Scuola Medica Salernitana”, University of Salerno, Salerno, Italy; Department of Psychiatry, University of Campania “Luigi Vanvitelli”, Naples, Italy; National Institute of Mental Health, Klecany, Czech Republic; Department of Psychiatry & Department of Child and Adolescent Psychiatry, Nagoya University Graduate School of Medicine, Nagoya, Japan; Montreal Neurological Institute and Hospital, McGill University, Montreal, Canada; Department of Psychiatry, Dokkyo Medical University School of Medicine, Mibu, Tochigi, Japan; Bipolar Center Wiener Neustadt, Sigmund Freud University, Medical Faculty, Vienna, Austria; Department of Clinical Psychiatry and Psychotherapy, Brandenburg Medical School, Brandenburg, Germany; Department of Psychiatry, National Institute of Mental Health and Neuroscience (NIMHANS), Bangalore-560029, India; Department of Genetic Epidemiology in Psychiatry, Central Institute of Mental Health, Medical Faculty Mannheim, University of Heidelberg, Mannheim, Germany; School of Psychiatry, University of New South Wales, and Black Dog Institute, Sydney, Australia; Department of Mental Health, Johns Hopkins Bloomberg School of Public Health, Baltimore, MD, United States; Department of Psychiatry and Behavioral Sciences, Norton College of Medicine, SUNY Upstate Medical University, Syracuse, New York, USA; Department of Psychiatry and Psychotherapy, University Medical Center (UMG), Georg-August University Göttingen, Göttingen, Germany; Department of Psychiatry and Psychotherapy, University of Münster, Münster, Germany; Department of Psychiatry, Melbourne Medical School, University of Melbourne, Parkville, Victoria, Australia; The Florey Institute of Neuroscience and Mental Health, The University of Melbourne, Parkville, VIC, Australia; Northern Adelaide Local Health Network, Mental Health Services, Adelaide, SA, Australia

**Keywords:** Lithium, polygenic score, Pharmacogenomics, Bipolar disorder, Psychiatry

## Abstract

**Background:** Polygenic scores (PGSs) hold the potential to identify patients who respond favourably to specific psychiatric treatments. However, their biological interpretations remain unclear. In this study, we developed pathway-specific PGSs (PS_PGS_) for lithium response and assessed their association with clinical lithium response in patients with bipolar disorder (BD).

**Methods:** Using sets of genes involved in pathways affected by lithium, we developed nine PS_PGSs_ and evaluated their associations with lithium response in the International Consortium on Lithium Genetics cohort (ConLi^+^Gen: N = 2367), validated in the combined PsyCourse (N = 105) and BipoLife (N = 102) cohorts. Lithium responsiveness was assessed using the Retrospective Assessment of the Lithium Response Phenotype Scale (ALDA scale), for categorical outcome (good vs poor response) and continuous ALDA total score. Logistic and linear regressions, adjusting for age, sex, chip type, and the first four genetic principal components, were used to test associations, after multiple testing corrections (*p*<0.05).

**Results:** Response to lithium was associated with PS_PGS_ for acetylcholine, GABA, calcium channel signalling, mitochondria, circadian rhythm, and GSK pathways, R² ranging from 0.29% to 1.91%, with R² of 3.71% for the combined PS_PGS._ Associations for GABA_PGS_ and CIR_PGS_ were replicated. In decile-based stratified analysis, patients with the highest genetic loading (10^th^ decile) for acetylcholine pathway genetic variants were 3.03 times (95%CI: 1.95 – 4.69) more likely to have a good lithium response than the lowest decile (1^st^ decile).

**Conclusion:** PS_PGSs_ achieved predictive performance comparable with conventional genome-wide PGSs, with more biological interpretability and using a smaller list of genetic variants, facilitating further investigation into the interaction of variants and biological pathways underlying lithium response.

## Introduction

Over the past 15 years, there has been significant progress in the development of polygenic scores (PGSs). Key research areas have been centred around evaluating their potential for disease risk prediction, uncovering the genetic basis of complex diseases, clinical application for disease screening and drug selection through pharmacogenomics, as well as assessing their cross-population transferability (1-5).

Since the initial implementation of the polygenic theory for assessing the genetic risk of schizophrenia (SCZ) by the International Schizophrenia Consortium in 2009 (1), hundreds of PGSs have been developed and investigated for their association with the risk of common mental health disorders such as SCZ (6), major depressive disorder (MDD) (7), and bipolar disorder (BD) (8). In recent years, PGSs have emerged as a promising tool for understanding the collective influence of common single nucleotide polymorphisms (SNPs) on patients’ pharmacological treatment outcomes (9, 10). For instance, in patients with SCZ, the PGS for SCZ (PGS_SCZ_) was significantly associated with antipsychotic treatment outcomes, reported to explain 3.2% of the interindividual variability in treatment response (11). Other studies have shown that PGS_SCZ_ can explain 2.0% of the variance in treatment resistance schizophrenia (TRS) (12-15), ∼1% in antipsychotic-induced weight gain (16, 17), nearly 2% in clozapine-induced myocarditis (18) and 2.7% in prolonged hospitalization (19). Similarly, in patients with MDD, the genetic score for SCZ, MDD, BD, and neuroticism showed significant associations with antidepressant treatment response (20-23) and resistance (24, 25), although each of these scores explained less than 2% of the variability. Among patients with BD, lithium treatment response was associated with the PGSs for SCZ (26) MDD (27), attention deficit hyperactivity disorder (28), and lithium responsiveness (29). Combined analysis of SCZ and MDD PGSs with clinical variables resulted in a better prediction, accounting for approximately 14% of the variance in lithium treatment response (30), emphasizing the potential clinical relevance of algorithms that combine PGSs with clinical data. This result exceeds the accuracy of any PGS analyzed individually or in combination using standard measures, which at best explain up to 5.6% of the variance in psychopharmacotherapeutic outcomes, e.g. in resistance to clozapine (31). While PGS hold significance for research purposes and offer promising clinical implications for the future, their predictive performance remains limited for direct clinical translation (10). Thus, there is a need to employ novel methods to develop PGSs with better predictive capabilities and to refine existing scores for increased precision.

In this context, newly proposed approaches such as biology-informed polygenic modelling have been evaluated for various traits (32-34). This PGS approach leverages genetic variants based on their relationship to molecular pathways that are linked to the phenotype of interest, thereby enhancing their predictive power and relevance to pharmacogenomics or disease screening (35). For example, an insulin receptor-based PGS targeting the striatum and prefrontal cortex predicted impulsivity and cognitive abilities in children, as well as addiction and dementia risk in adults (32). Similarly, PGS composed of variants associated with nervous system development and neuron differentiation explained 6.9% of the variance in liability to psychosis, in a sample of patients with DSM-IV diagnoses of SCZ or psychosis-related disorders determined by structural clinical interview. This result surpasses the 3.7% of the variance explained by conventional PGS_SCZ_ using genome-wide variants (34). Thus, restricting PGS to genetic variants within biological pathways known to be associated with lithium response, may reduce “noise” from variants with spurious associations and increase the power of polygenic models, while explicitly building our mechanistic understanding (36). Furthermore, the biology-informed polygenic approach may facilitate the effort to identify new treatment targets (33, 37).

In our recent study, the cholinergic and glutamatergic pathways were enriched in the genome-wide association study of lithium response and a statistically significant association was found between high genetic loading for lithium-responsive variants and lithium responsiveness phenotypes (29). Expanding this concept, we undertook a literature review of the wider current understanding of biological pathways and processes impacted by lithium exposure, and then developed pathway-specific PGS (PS_PGS_) for lithium response. We hypothesized that these PS_PGS_ would predict lithium response in patients with BD.

## Methods and materials

### Study sample characteristics

The target data for this study was obtained from the International Consortium on Lithium Genetics (ConLi^+^Gen) (http://www.conligen.org/), a global initiative established to investigate the genetic underpinnings of lithium treatment response in patients with BD. The discovery and target sample included only patients of European ancestry (N= 2367) who received lithium and followed up for at least six months (38). The number of participants in each country is described in our previous study (28).

To replicate the findings from ConLi^+^Gen, we utilized combined data from two German cohort studies: the pathomechanisms and signature in the longitudinal course of psychosis study (PsyCourse, N=105) (39, 40) and BipoLife cohort (N= 102) (41). A detailed sample selection procedure for the replication cohorts is included in Supplementary material 1.

*Insert supplementary material 1 here*

### Target outcome measure

For both target and replication cohorts, the validated retrospective criteria of long-term treatment response in research subjects, known as the ALDA scale, was used to assess patient’s response to lithium treatment (42, 43). This scale quantifies the degree of improvement during lithium response expressed as a composite measure of change in frequency and severity of mood symptoms (A score). This score is weighted with five determinant factors of symptom improvement (B scale). These determinant factors are the number (B1) and the frequency (B2) of episodes before/off the treatment, the duration of the treatment (B3), and the compliance and use of additional medication during the periods of stability. The total Alda score was calculated by subtracting the B score from the A score. The target outcome — lithium response was defined in categorical (good response vs. poor response) and continuous outcomes. For the categorical outcome, patients having a total score of 7 or higher were classified as “good responders,” and a score of less than seven were classified as “poor responders” (44). The total Alda score was used as a continuous lithium response measure, after excluding patients with B scores greater than four or who had missing data. Negative scores were recalibrated as 0. This algorithm has been used in previous studies (26, 27, 30, 45, 46) and described in detail elsewhere (44).

### Genotyping, quality control, and imputation procedures for ConLi^+^Gen sample

DNA was extracted from peripheral blood samples collected at 22 participating sites, and samples were genotyped using either Affymetrix or Illumina SNP arrays (44). Prior to imputation, quality control (QC) procedures were implemented on the genotype data using PLINK version 1.9 (47). SNPs with a poor genotyping rate (< 95%), strand ambiguity (A/T and C/G SNPs), a minor allele frequency (MAF) less than 10%, and SNPs deviated from Hardy-Weinberg Equilibrium (HWE) (*p* < 10^-6^) were removed. Individuals with sex inconsistencies between the documented and genotype-derived sex and genetically related were also excluded. The genotypic and quality control details of the ConLi^+^Gen cohort are available elsewhere (44).

The genotype data passing QC were imputed in the Michigan server separately for each genotyping platform using the Haplotype Reference Consortium (HRC) reference panel comprising broadly European haplotypes at 39,235,157 SNPs (48). For each cohort, imputation quality procedures were implemented and excluded SNPs of low-frequency (MAF) < 1%) and low-quality (imputation quality score R^2^ < 0.6). Then, genotype calls for the filtered SNPs were derived and merged using PLINK from the imputed dosage score (47). The genotyping, quality control, and imputation procedures for the replication cohorts are found in supplementary material 1.

### Developing biologically informed polygenic scores

***Step one:*** *Identify biological pathways (targets) of lithium*.

To develop a PS_PGS_, we first conducted a narrative review to identify biological pathways or processes potentially modulated by lithium, followed by extraction of candidate genes and genetic variants involved in these pathways (*Supplementary materials 1).* Our review identified pathways including acetylcholine, Gamma-aminobutyric acid (GABA), glutamate, dopamine, calcium channels, mitochondria, circadian rhythm, Glycogen Synthase Kinase (GSK) and N-methyl-D-aspartate (NMDA) as potential targets for lithium in treating patients with BD.

***Step two:** Map genes and single nucleotide polymorphism for each biological pathway*

Using three existing databases, specifically, gene set enrichment Analysis (GSEA) GSEA (gsea-msigdb.org), HUGO gene Nomenclature committee (HGNC) Home | HUGO Gene Nomenclature Committee (genenames.org) and Kyoto encyclopedia of genes and genomics (KEGG) https://www.genome.jp/kegg/, candidate genes associated with the potential biological targets (pathways) of lithium (step 1) were extracted using the relevant/pathway name as a search term. For instance, the term “mitochondria” was used to select relevant gene sets within these databases. In total, 942 genes in nine potential biological pathways were identified, i.e., acetylcholine (164), GABA (76), glutamate (92), dopamine (155), calcium channel (134), mitochondria (163), circadian rhythm (129), GSK (18) and NMDA (11). These gene sets are selected based on direct association with lithium pharmacokinetics; they may not capture all possible gene sets in each pathway, and there are few overlapping genes across pathways *(Supplementary material 1)*. Then, SNPs located on these genes for each pathway were annotated using the Genome Reference Consortium Human Build 37 (GRCh37/hg19) reference panel and Multi-marker Analysis of GenoMic Annotation (MAGMA) software https://ctg.cncr.nl/software/magma (49), with –*annotate window = 100, 20* (100kb upstream and 20kb downstream window). With this process, we mapped 38,933 SNPs i.e., acetylcholine (6247), GABA (2994), glutamate (3840), dopamine (5794), calcium channel (4236), mitochondria (7801), circadian rhythm (6673), GSK (707) and NMDA (641). The list of genes and SNPs is provided in the *supplementary material 2*.

*Insert supplementary material 2 here*

**Step three:** *Develop pathway-specific polygenic scores*.

In the ConLi+Gen cohort, participants were selected from 13 countries, and we accounted the potential genetic variation by participants’ country of origin – considering potential geographical differences in ethnicity, we implemented a widely accepted leave-one-country-out (LOC) procedure (50, 51) where PGSs were calculated for participants of one country at a time (target sample) using GWAS summary statistics from the remaining 12 countries (discovery sample). We generated 26 GWAS summary statistics for categorical and continuous lithium responses. Each GWAS was performed using PLINK adjusting for regression models adjusted for age, sex, chip type, and the first four principal components (PCs). Each of the PS_PGSs_ was computed using PGS-CS method, which incorporates continuous shrinkage priors on effect sizes and accounting for linkage disequilibrium (LD) among SNPs (52). In the replication analysis, a summary GWAS data from the full ConLi^+^Gen was used as discovery sample (29) to compute PS_PGS_ in the PsyCourse (39, 40) and BipoLife samples (41). Further details on the development of PS_PGS_ are provided in the supplementary material 1.

### Step four: Association analysis

Finally, the associations of each of the PS_PGSs_ and lithium treatment response was evaluated using linear regression for the continuous outcome and logistic regression for the categorical outcomes. Each association analysis was adjusted for age, sex, chip type and the first four PCs. A statistically significant association was determined at *p* < 0.05 after multiple testing using the Benjamini-Hochberg Procedure (BH) (53).

To evaluate the combined effect of multiple PS_PGSs_ on lithium response, we performed a multivariate regression modeling considering only PS_PGSs_ that showed a significant association with lithium response in the univariate model. The performance of this combined PS_PGS_ model was compared with the conventional PGS model that uses whole genome genetic variants in predicting lithium (54). Stratified analysis was conducted by dividing the ConLi^+^Gen sample into deciles, ranging from the lowest to the highest polygenic loading for each PS_PGSs_. High PGS indicates patients with a higher polygenic load for the specific pathway, while low PGS indicates the opposite. The proportion of phenotypic variance explained (R^2^) by each PS_PGSs_ was calculated as the difference in R^2^ between the model fit with specific PGSs and covariates, and the model with only covariates. Mc Fadden pseudo R^2^ was calculated for categorical lithium response (55) and the reported observed R^2^ was transformed to a liability scale (56) considering a population prevalence of lithium responsiveness of 30% (44, 57) and the proportion of lithium responders to nonresponders in the target and replication cohorts.

**Figure 1:**
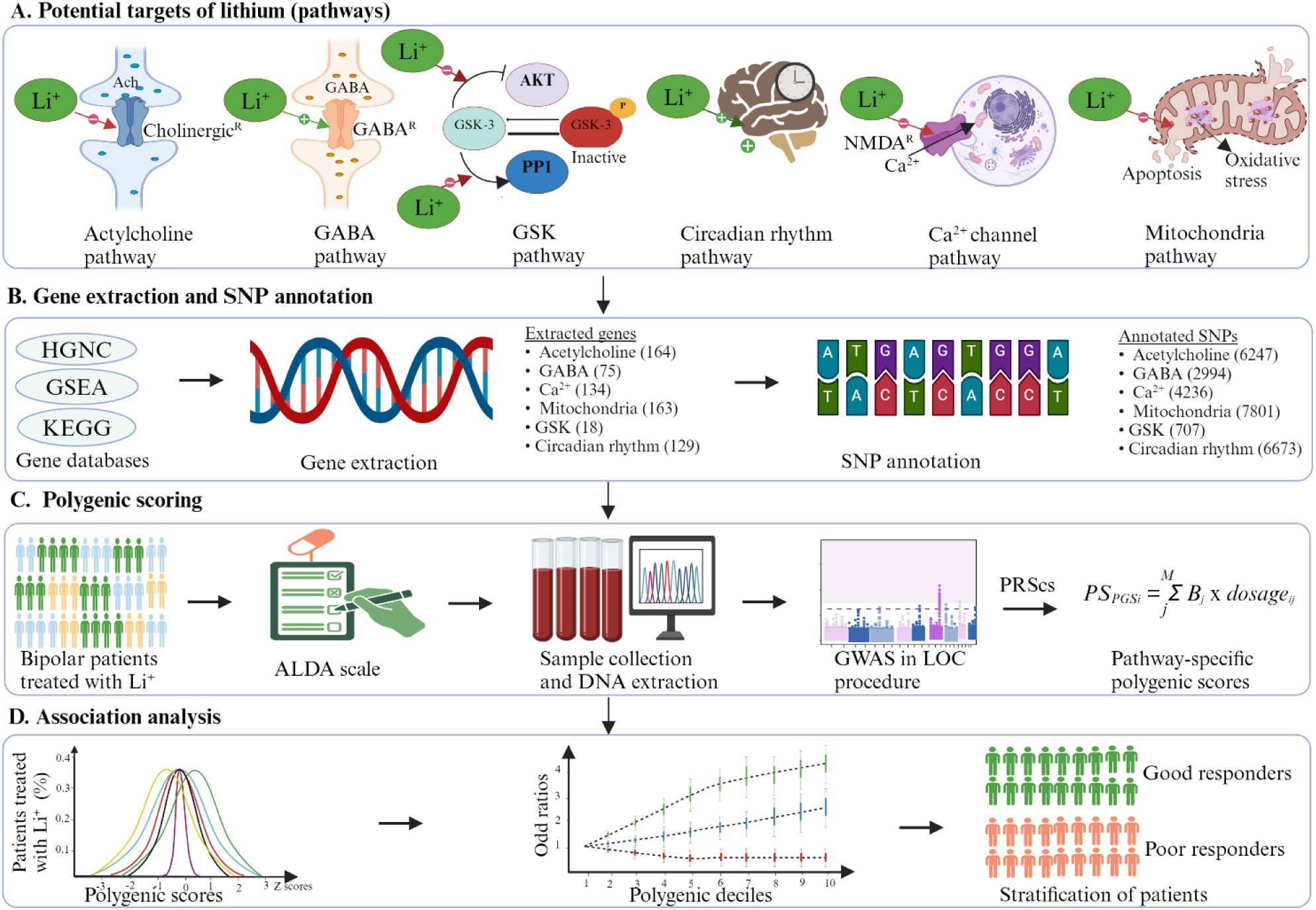
The schematic presentation of the procedures of pathway selection and detailed steps of the data analysis process. *Abbreviations*: ConLi^+^Gen = The International Consortium on Lithium Genetics; SD = Standard deviation; ALDA = Retrospective Criteria of Long-Term Treatment Response in Research Subjects with Bipolar Disorder

## Results

### Description of study participants

Our discovery analysis from ConLi^+^Gen included data from 2367 patients with BD treated with lithium for at least six months. Nearly 60% of the participants were females; the mean age (SD) was 47.53 (13.73) years. Six hundred sixty (27.9%) patients had a good response to lithium treatment (defined as ALDA score ≥7) and the mean (SD) ALDA total score was 4.12 (3.15). Among the replication cohort participants, 48.0% of the BipoLife and 40% of PsyCourse were females. About 28% of patients in the Bipolife cohort and 23% of patients in the PsyCourse cohort had good response to lithium treatment (Table 1).

**Table 1:** Characterises of the study cohorts and participants.

| Cohort | N | Sex<br>(female) | Age (SD) | ALDA score,<br>Mean (SD) | Good response to<br>lithium, N (%) |
| --- | --- | --- | --- | --- | --- |
| ConLi<sup>+</sup>Gen | 2367 | 1369 | 47.53(13.73) | 4.12(3.15) | 660(27.88) |
| BipoLife | 102 | 49 | 49.87(13.62) | 4.52(2.93) | 29(28.43) |
| PsyCourse | 105 | 42 | 46.91(12.64) | 3.80(2.87) | 24(22.86) |

*Abbreviations:* ConLi<sup>+</sup>Gen = The International Consortium on Lithium Genetics; SD = Standard deviation; ALDA = Retrospective Criteria of Long-Term Treatment Response in Research Subjects with Bipolar Disorder

### Associations of pathway-specific polygenic scores for lithium response with lithium treatment response

**Figure 2:**
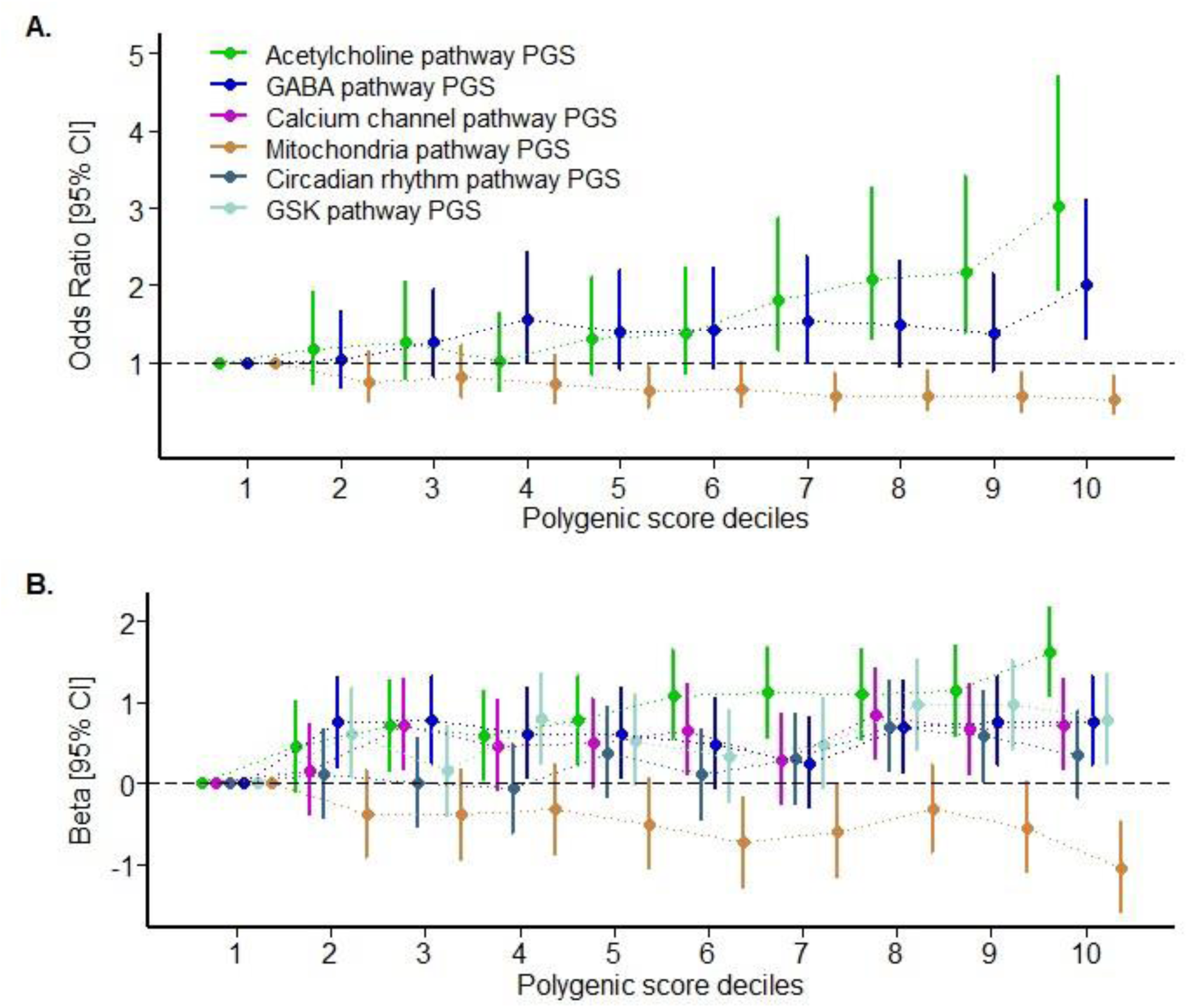
Trends in the odds ratios for favourable lithium treatment response (with the categorical (A) and continuous (B) outcomes in patients with bipolar disorder, comparing those with a high pathway-specific PGS, deciles (2^nd^ to 10^th^) with those patients with the lowest genetic score (decile 1^st^) (n = 2367) for the pathways that had a significant association after multiple testing. *Legends:* The dot points and error bars represent the odd ratio and 95% confidence interval for the respective polygenic deciles. The polygenic score deciles that crossed the odd ratio one on the y-axis are not statistically significant. *Abbreviation:* GABA = Gamma-aminobutyric acid

Bipolar patients with higher PGS for acetylcholine genetic variants (ACh_PGS_) were more likely to have good lithium treatment response than the lower; aOR = 1.34 (95%CI: 1.22 – 1.49), *p* = 3.54x10^-8^; R^2^ = 1.91%) in categorical and 0.38 (95%CI: 0.26 – 0.51, *p* = 3.24x10^-9^; R^2^ = 1.56%) in continuous outcomes. In the stratified analysis, patients with the highest PGS decile in ACh_PGS_ were 3.03 times (95%CI: 1.95 – 4.69) more likely to have a good lithium response than their counterparts. Similarly, higher GABA pathway PGS (GABA_PGS_) was associated with a greater likelihood of good response to lithium — both in the categorical (*p* = 9.14x10^-3^; R^2^ = 0.34%) and continuous outcomes (*p* = 0.03; R^2^ = 0.30%), with a 2.01-fold increase in the highest decile (95%CI: 1.30 – 3.09). Calcium channel pathway PGS (Ca^2+^) was significant with continuous lithium response (*p* = 0.01; R^2^ = 0.3%) but not with categorical outcome (*p* = 0.11). GSK pathway PGS (GSK_PGS_) and Circadian Rhythm Pathway PGS (CIR_PGS_) were positively associated with good lithium response — with continuous (*p* = 1.84x10^-4^; R^2^ = 0.6%) and (*p* = 0.01; R^2^ = 0.2%) respectively, but not with categorical outcome. In contrast, the increased genetic variance within mitochondria genes was associated with poorer lithium response — with categorical (*p* = 7.42x10^-4^; R^2^ = 1.05%) and continuous outcomes (*p* = 1.02x10^-4^; R^2^ = 1.16%) in mitochondria pathway PGS (MITO_PGS_). With a decreasing trend across deciles, patients with the highest MITO_PGS_ had 51% lower odds of responding to lithium than the lowest decile; aOR = 0.52 (95%CI: 0.33 – 80). The remaining PS_PGSs_ were not significant. The full stratified analysis is presented in Supplementary Material 1.

### Combined modelling of pathway-specific PGS

The combined modelling of ACh_PGS_, GABA_PGS_, Ca^2+^, MITO_PGS,_ GSK_PGS_, and CIR_PGS_ collectively explained 3.71% of the variance in categorical and 3.18% in continuous lithium treatment responses. These results were higher than the variance explained by each of the PS_PGSs_ (Table 2) and comparable with the conventional genome-wide PGS *(Supplementary material 1)*.

**Table 2:**
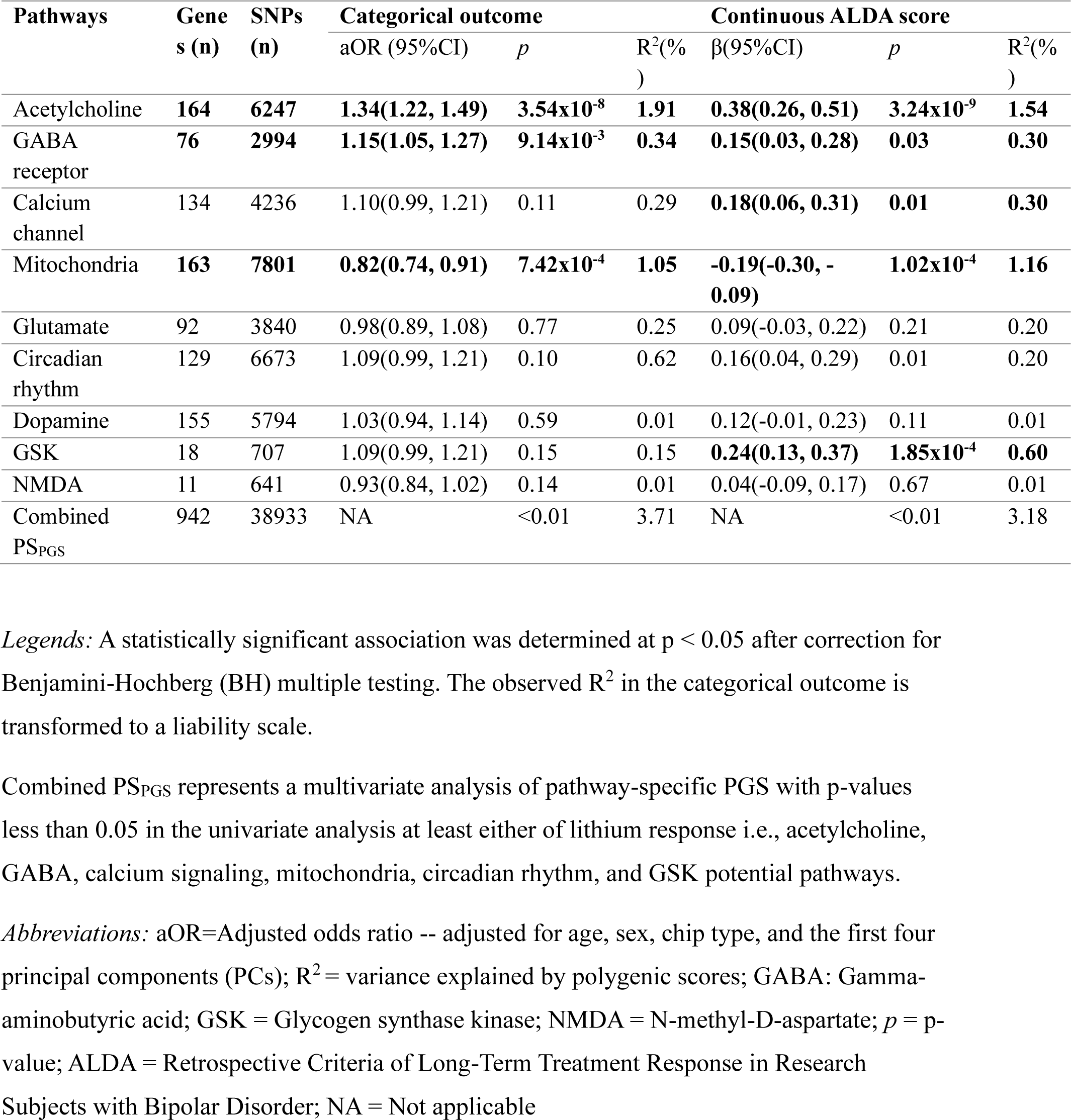
The association of pathway-specific-PGSs and lithium treatment response in patients with bipolar disorder.

### Replication analysis

Using the combined datasets from PsyCourse and BipoLife, we found a statistically significant association between GABA_PGS_ and lithium treatment response, both in the categorical (*p* < 0.01; R^2^ = 2.74%) and continuous outcomes (*p* = 0.01; R^2^ = 3.30%), replicating the findings from the discovery analysis. The results showed that bipolar patients who had a higher GABA_PGS_ were 1.53 times (95%CI: 1.09 – 2.14; in the categorical) and 0.57 times (95%CI: 0.18 – 0. 96; in continuous outcomes) more likely to have a good response to lithium compared with those with the lowest GABA_PGS_. The CIR_PGS_ was also significantly associated with continuous lithium response (*p* = 0.01, R^2^ = 1.38), and with each one-unit increase in the CIR_PGS_, there is an associated increase in the likelihood of a good response to lithium by 0.40 (*β* = 0.40, 95% CI: 0.01–0.79). The association results of PGS for other potential biological pathways were not replicated. The full replication analysis results are available in *Table 3*.

**Table 3:** The association of pathway-specific-PGSs and lithium treatment response among bipolar patients treated with lithium in replication cohorts (combined PsyCourse and Bipolife cohorts)

| Pathways | Gene<br>s (n) | SNPs<br>(n) | Categorical outcome |  |  | Continuous outcome |  |  |
| --- | --- | --- | --- | --- | --- | --- | --- | --- |
| | | | aOR (95%CI) | R<sup>2</sup>(%) | *p* | $\beta$ (95%CI) | *p* | R<sup>2</sup>(%) |
| Acetylcholine | 164 | 6247 | 1.21(0.88, 1.67) | 0.95 | 0.09 | 0.24(-0.15, 0.64) | 0.22 | 0.22 |
| GABA | **76** | **2994** | **1.53(1.09, 2.14)** | **2.74** | **0.01** | **0.57(0.18, 0.96)** | **0.01** | **3.31** |
| Calcium<br>channel | 134 | 4236 | 0.96(0.69, 1.33) | 0.05 | 0.79 | 0.25(-0.14, 0.65) | 0.21 | 0.30 |
| Mitochondria | 163 | 7801 | 0.96(0.70, 1.33) | 0.03 | 0.83 | -0.13(-0.52, 0.27) | 0.51 | 0.01 |
| Circadian<br>rhythm | 92 | 3840 | 0.99(0.72, 1.37) | 0.02 | 0.94 | **0.40(0.01, 0.79)** | **0.04** | **1.38** |
| Glutamate | 129 | 6673 | 0.98(0.71, 1.35) | 0.02 | 0.88 | 0.29(-0.10, 0.68) | 0.15 | 0.54 |
| Dopamine | 155 | 5794 | 1.07(0.78, 1.47) | 0.13 | 0.67 | -0.20(-0.60, 0.19) | 0.31 | 0.01 |
| GSK | 18 | 707 | 1.13(0.82, 1.58) | 0.33 | 0.44 | 0.01(-0.39, -0.40) | 0.96 | 0.01 |
| NMDA | 11 | 641 | 1.13(0.77, 1.69) | 0.03 | 0.81 | 0.13(-0.26, 0.52) | 0.52 | 0.01 |

*Legends:* A statistically significant association was determined at $p < 0.05$ after correction for Benjamini-Hochberg (BH) multiple testing. The analysis models were adjusted for age and sex. The observed $R^2$ in the categorical outcome is transformed to a liability scale.
*Abbreviations:* aOR=Adjusted odds ratio; $R^2$ = variance explained by polygenic scores; GABA: Gamma-aminobutyric acid; GSK = Glycogen synthase kinase; NMDA = N-methyl-D-aspartate; $p$ = p-value; ALDA = Retrospective Criteria of Long-Term Treatment Response in Research Subjects with Bipolar Disorder

## Discussion

For the first time, we developed biologically informative PS_PGSs_ in well-characterized datasets and evaluated their association with lithium treatment response in patients with BD. Building upon a previous study (29) that employed the conventional PGS approach (utilizing genome-wide genetic variants), the current analysis targeted genetic variants mapped within acetylcholine-, GABA-, calcium channel-, mitochondria-, glutamate-, circadian rhythm-, dopamine-, NMDA and GSK pathways that are characterized as potential pharmacological targets for lithium (58, 59). We found that BD patients with higher genetic loading of variants within the acetylcholine-, GABA-, calcium channel-, GSK and circadian rhythm pathways were more likely to respond to lithium treatment. In contrast, individuals with higher loading for genetic variants in the mitochondria pathway were less likely to respond to lithium. Our stratified analysis showed that patients with the high genetic loading for acetylcholine-, GABA- and calcium channel pathways variants (in the 10^th^ decile) have good lithium treatment response compared to those with the lowest genetic loading (1^st^), with an increasing trend of lithium treatment responsiveness across 1^st^ decile to 10^th^ decile. The trend was reversed in the mitochondria pathway.

A combined PS_PGS_ explained 3.71% of the phenotypic variance variability in categorical and 3.18% in the continuous lithium treatment response, comparable to the predictive power of the conventional polygenic model developed using genome-wide variants (29). The PS_PGS_ approach incorporates biological information by restricting SNPs in each pathway-specific PGS to those within genes related to a single pathway or biological phenotype of interest (e.g. circadian rhythm, mitochondrial function), allowing for targeted testing of associations between pathway-specific genetic variants and phenotypic outcomes (60). Modeling individual variations in lithium-related pathways may capture significant enrichment of genetic variants at pathway-level, highlighting PS_PGS_ as a more biologically interpretable, higher-resolution alternative to genome-wide PGS (60). Moreover, PS_PGS_ could facilitate the development of novel drugs through drug repurposing that targets the selected genes in each pathway (61). Pathway-specific PGS approach also prioritizes variants that may contribute to a higher heritability estimate and may detect enriched and functionally relevant GWAS signals in contrast to reliance on statistically associated variants (62-65). These features minimize noise variants and leverage the genetic variation across multiple potential biological pathways, which has the potential to achieve better clinical utility of PGSs. Combining these scores with clinical data may further improve variance explained to improve clinical utility (30).

In terms of the directions of associations with lithium treatment response, ACh_PGS,_ GABA_PGS_, Ca^2+^, CIR and GSK were *positively* associated. For instance, in the decile-based stratified analysis, patients with the highest loading for acetylcholine pathway genetic variants (10^th^ ACh_PGS_ decile) were 3.03 times (95%CI: 1.95 – 4.69) more likely to have a good lithium response compared to those with the lowest genetic loading (1^st^ decile). These positive associations between ACh_PGS_, GABA_PGS_, and lithium treatment response are consistent with evidence suggesting that lithium acts to correct deficits in ACh and GABA neurotransmission (58, 59, 66, 67). A similar association between lithium treatment response and Ca^2+^ is consistent with evidence of disruption of cellular calcium concentrations in patients with BD (68). Lithium is known to attenuate calcium release and regulate intracellular calcium levels in hippocampal neurons, thereby reducing excitotoxicity (69). Regarding the circadian rhythm pathway, lithium is widely recognized for its efficacy in improving sleep rhythm by increasing amplitude and slowing rhythm cycles (70, 71). Good lithium responders show higher amplitude sleep cycle (72) and lithium has been found to correct rhythm abnormalities in patients with BD (73, 74). The positive association between GSK_PGS_ and lithium treatment response align with previously reported functional enrichment of the PI3K-Akt signalling pathway, which involves GSK3β and associated with response to lithium (75). From the above evidence, a higher genetic loading appears to be related to better treatability.

On the other hand, the increased genetic variance within mitochondrial genes was associated with poorer lithium treatment response. Evidence suggests that BD patients may experience reductions in mitochondrial enzyme levels and overall mitochondrial health, resulting in reduced bioenergetic capacity (76). Mitochondrial gene expression tends to be lower in the post-mortem brains of patients with BD and is rescued specifically in lithium responders via a number of potential mechanisms including expression of electron transport chain proteins, second messenger systems such as PKA, PKC and in intracellular potassium and calcium regulation (76-78). Studies in IPSC-derived neuron culture suggest that lithium may act to correct hyperexcitability via these mechanisms (76). The significant association between Mito_PGS_ and lithium treatment response underscores the centrality of mitochondrial health in lithium’s mechanism of action. The relationship between our negative association of these genes with lithium response and protein expression networks needs further exploration.

Pathway-specific PGSs have been employed in several studies to enhance risk stratification and improve treatment efficacy (79-82). For example, Grama et al. investigated whether behavior- and neuronal-related gene sets, previously implicated in SCZ, were associated with subcortical volumes (79). They found that PGS derived from an abnormal behaviour gene set was associated with right thalamic volume, and this association was robust across p-value thresholds, unlike the finding from a genome-wide approach (79). Warren and colleagues also studied the relationship between the genome-wide PGS for SCZ and the neurotransmitter PGSs (Glutamate, GABA, Dopamine, and Serotonin) with psychotic disorder presentation (80). In this study, there was no significant association between individual symptom measures and SCZ_PGS_, while glutamate and GABA pathway PGSs were associated with psychosis case status, and dopamine pathway PGS was significantly associated with poorer global functioning in psychosis subjects (80). Other studies have shown that the PGS for oxidative stress pathway significantly differentiated individuals with early psychosis status from the controls (82), and dopamine pathway PGS has been implicated in the pathophysiology of schizophrenia (81).

Other examples of the development of PS_PGS_ in cardiovascular medicine have showcased their potential in personalizing treatment approaches. For instance, PGS for calcium signalling pathways along with phosphatidylinositol/inositol phosphate pathways was associated with hypertensive status, suggesting their regulation could be a potential pharmacological preventive target for hypertension (83). PGS tailored to pharmacodynamic pathways of angiotensin-converting enzyme (ACE) inhibitors (84) and β-blockers have respectively enhanced patient selection for ACE inhibitors and predicted mortality in heart failure (85). Similarly, a pharmacogenomic polygenic response score developed 31 genes associated with adenosine diphosphate-based platelet reactivity during clopidogrel treatment has been applied to predict major adverse cardiovascular events and cardiovascular death in patients with coronary artery disease treated with clopidogrel (86).

Drawing from the evidence presented in our study and the above studies, PS_PGSs_ hold promise for the future of precision psychiatry by refining patient treatment stratification and improving treatment efficacy through leveraging genetic information across specific pharmacological pathways. For lithium treatment this has multiple implications, the stratification of likely good and poor responders could reduce delays to effective treatment and associated burden. Serious side effects of long-term lithium therapy, including chronic renal failure and mortality due to toxicity (87), can also be minimized.

## Limitations

While our study provides novel and robust support for the use of PS_PGS_ methods, the results should be interpreted in conjunction with some limitations. *First,* since it is an observational study, it is challenging to draw definitive conclusions about causality in the observed associations. The ALDA scale used to define lithium response considers the impact of polypharmacy, adherence and longitudinal efficacy but may be affected by recall bias (88). *Second*, only participants of European ancestry were included as this study builds upon our previous research (29), and these results cannot be generalized to other ancestrally diverse populations. Expanding this work to include the Asian ConLi^+^Gen cohort would indeed be valuable, as it could reveal whether the findings replicate across different ancestries. *Third*, while our general strategy for pathway selection was based on the narrative review, the cholinergic and glutamatergic pathways were identified from our previous work (29), that utilized the same sample. The PGSs derived from significant pathways in the ConLi^+^Gen sample could likely show statistically significant association within this sample, emphasising the need for future replication in independent datasets. However, it is important to note that PsyCourse and BipoLife cohorts were not included in the original pathway analysis, and replicating the findings in this cohort minimizes potential bias. *Fourth,* we used relatively small cohorts for the replication analysis, and larger replication-independent cohorts are needed. The results for acetylcholine, calcium signalling and mitochondrial pathways were not replicated in our replication analysis. *Fifth*, our search was not exhaustive, and we may have excluded important biological pathways implicated in lithium pharmacology. *Sixth,* biological pathways are highly complex and their relationships, interactions, and direct links with function are not yet comprehensively mapped, thus limiting the variance explained in medication response.

## Conclusion

By focusing on biologically relevant genetic variants, PS_PGS_ for lithium response has shown predictive capabilities comparable to the conventional genome-wide PGSs, but with the advantage of using fewer number of SNPs and providing biological interpretability While the variance in lithium treatment response explained by these models is still small, at best 3.71%, future models may include an expanded range of lithium-specific pathways to improve accuracy, reaching an effect size relevant to stratifying individuals by genetic risk for personalization of lithium treatment. Our study invites further investigation of how proteins including acetylcholine, GABA, calcium signalling, mitochondria, GSK, glutamate and circadian rhythm pathways interact at the molecular level to define lithium treatment response. Replication in larger cohorts, including non-European ancestry is required to establish pathway-specific PGS’s clinical utility.

## Data availability

**All data used in this analysis is available to ConLi_+_Gen members.**

See http://www.conligen.org/ for more information.

**Software**

Here are the software: **PRS-cs,** see, https://github.com/getian107/PRScs; for **MAGMA,** see https://ctg.cncr.nl/software/magma; for **PLINK 2,** see https://zzz.bwh.harvard.edu/plink/tutorial.shtml. The custom codes used in this study will be made available upon reasonable request to the corresponding author.

## Acknowledgment

The authors are grateful to all patients who participated in the study, and we appreciate the contributions of clinicians, scientists, research assistants, and study staff who helped in the patient recruitment, data collection, and biological sample preparation of the studies. We are also indebted to the members of the ConLi^+^Gen Scientific Advisory Board (http://www.ConLi+Gen.org/) for critical input over the course of the project.

Angel’s pilot work, as part of her honours thesis, provided the foundation for designing this project, and we gratefully acknowledge her contribution. The analysis of this study was carried out using the high-performance computational (HPC) capabilities of the University of Adelaide’s Phoenix Supercomputer https://www.adelaide.edu.au/phoenix/.

## Author contributions

AT Amare conceived and designed the project and secured a fellowship to lead the study. NT Sharew developed the research proposal, conducted the statistical analysis, interpreted the findings, and drafted the manuscript. AT Amare, SR Clark, and KO Schubert provided supervision and critically reviewed the data analysis steps and manuscript draft. All authors contributed genetic and clinical data, provided feedback, and made significant intellectual contributions to the manuscript.

