## Supplementary Material 1 for "Pathway-Specific Polygenic Scores for Lithium Response for Predicting Clinical Lithium Treatment Response in Patients with Bipolar Disorder"

### **Selecting potential biological pathways of lithium in the treatment of patients with bipolar disorder: Narrative review**

**Methods:**

The narrative was performed in the PubMed database from May 09, 2014 to May 09, 2024 using the search string: ((lithium[Title/Abstract]) OR (lithium salt[Title/Abstract]) AND (“Bipolar Disorder”[Mesh] OR Bipolar disorder[Title/Abstract] OR Bipolar dis*[Title/Abstract]) AND (“Pharmacology"[Mesh] OR "pharmacology" [Subheading] OR mechanism of action[Title/Abstract] OR mechanism*[Title/Abstract])). Following title and abstract screening, 1171 studies were identified, with 38 studies undergoing full-text review. Our search strategy included original research and systematic reviews exploring the mechanism action of lithium in the treatment of patients with bipolar disorder (BD) while excluding conference abstracts, case reports, editorials and notes. Publications other than the English language were also excluded.

Thirteen studies met inclusion/ exclusion criteria and were analyzed to select the potential biological pathways in lithium treatment. The details of the search strategy and results are summarized in table 1 below.

**Supplementary Table 1:** Complete search strategy for the narrative review

| Concepts | Search strings (PubMed) | PubMed |
| --- | --- | --- |
| Lithium (#1) | (lithium[Title/Abstract]) OR (lithium salt[Title/Abstract]) | 66,015 |
| Bipolar disorder (#2) | "Bipolar Disorder"[Mesh] OR Bipolar disorder[Title/Abstract] OR Bipolar dis*[Title/Abstract] | 59,820 |
| Mechanism of action (#3) | “Pharmacology"[Mesh] OR "pharmacology" [Subheading] OR mechanism of action[Title/Abstract] OR mechanism*[Title/Abstract] | 9,000,514 |
|  | #1 AND # 2 AND #3 | 4801 |
|  | Studies in the last 10 years | 1171 |
|  | Studies included after title and abstract search | 38 |
|  | Records identified from citation searching | 1 |
|  | Studies included in the final analysis after full-text review | 13 |

From these studies, we identified lithium response related biological phenotypes including those associated with neurotransmission, biological second messenger systems, and clinical features linked to treatment response in BD patients including: acetylcholine, Gamma-aminobutyric acid (GABA), calcium signalling, mitochondria, glutamate, circadian rhythm, dopamine, N-methyl-D-aspartate (NMDA) and Glycogen synthase kinase (GSK) (see table 2). To support subsequent search for related genes we then describe the features of biological pathways related to each phenotype in our review.

**Supplementary Table 2:** Identified potential biological pathways in the treatment of lithium

| Author (year) | PubMed ID | Key finding (Reported potential pathways) |
| --- | --- | --- |
| Malhi & Outhred (2016) (1) | 27638546 | Acetylcholine, GABA, Calcium signalling and Mitochondria, Dopamine, glutamate, NMDA, GSK and circadian rhythm |
| Malhi et al (2023) (2) | 23371914 | Acetylcholine, GABA, Calcium signalling and Mitochondria, Dopamine, glutamate, NMDA, GSK and circadian rhythm |
| Van Enkhuizen et al (2015) (3) | 26141192 | Acetylcholine |
| Amare et al (2023) (4) | 37433967 | Acetylcholine |
| McCarthy et al (2016) (5) | 26476274 | Calcium Channel Signaling |
| D’onofrio et al (2016) (6) | 27033453 | Calcium Channel Signaling |
| Mertens et al (2015) (7) | 26524527 | Calcium Channel Signaling and mitochondria |
| Stacey et al (2018) (8) | 30185780 | Mitochondria |
| Osete et al (2021) (9) | 34075196 | Mitochondria |
| Lundberg et al (2020) (10) | 33132941 | Mitochondria |
| De Sousa et al (2015) (11) | 24961563 | Mitochondria |
| Geoffroy et al (2018) (12) | 28095742 | Circadian rhythm |
| Geoffroy et al (2016) (13) | 27324142 | Circadian rhythm |

*Abbreviations:* GABA = Gamma-aminobutyric acid; GSK = Glycogen synthase kinase (GSK); NMDA = N-methyl-D-aspartate;

The mechanisms of lithium response are many and varied. Here we perform a mini-narrative review of potential biological pathways associated with lithium treatment response in patients with BD.

Acetylcholine is a neurotransmitter that binds to two main receptor subtypes - muscarinic and nicotinic (14). Muscarinic receptor activation stimulates a cascade of second messenger interactions, including G protein and phospholipase, inducing calcium release. Through G proteins, muscarinic receptors also inhibit adenylate cyclase activity and reduce the formation of cyclic adenosine monophosphate (cAMP) (15). Chronic lithium intake increases acetylcholine synthesis and choline uptake (16). Additionally in culture, lithium enhances the expression of muscarinic M3 receptors while decreasing M2 receptor levels (17). In humans, acetylcholine levels are elevated across brain regions in depressed patients and lithium treatment has been shown to normalise acetylcholine-induced behavioural despair (3, 18). These findings support that the acetylcholine and particularly muscarinic receptor related gens may be relevant in predicting the response to lithium treatment.

GABA is an inhibitory neurotransmitter that plays a key role in mood stabilization, and patients with BD have low GABA-ergic neurotransmission (19, 20). Low levels of GABA in plasma and cerebrospinal fluid in patients with BD increases excitatory neurotransmission, leading to excitatory toxicity and potentially neuronal apoptosis (21, 22). Lithium promotes the release of neuroprotective proteins and reduces neuronal excitation by increasing the level of GABA (23, 24). These findings suggest that the GABA pathway and its genetic variations may be relevant in predicting the response to lithium treatment in BD patients.

Glutamate is an excitatory neurotransmitter (25), and lithium increases active regulatory elements, such as the enhancers and promoters of genes involved in glutamate neurotransmission at the cellular level (26). Lithium treatment has also been linked with decreased phosphorylation of subunits of N-methyl-D-aspartate (NMDA) receptors in the prefrontal cortex and ventral striatum, and associated reduction in glutamate levels (27). Additionally, variants of the glutamate decarboxylase-like protein (*GADL1*) gene have been shown to predict lithium treatment response BD patients (28). These findings denote that the glutamate pathway and its genetic variations may be relevant in predicting the response to lithium treatment BD patients.

N-methyl-D-aspartate receptors have been implicated in the pathophysiology of BD(29) Lithium inhibits the NMDA receptors which subsequently reduces the level of glutamate and NMDA receptor-mediated calcium influx (1). The NMDA activation is also influenced by other neurotransmitters that are altered by lithium. For instance, lithium has been shown to reduce dopamine activity which increases NMDA receptors (30). Thus, investigating NMDA receptors and their genetic variation might be relevant in predicting lithium treatment response.

Dopaminergic dysfunction has been identified as a potential mechanism underlying the pathogenesis of BD (31, 32). Post-mortem studies suggest the upregulation of dopamine receptors in patients with BD (33, 34). Lithium induces second messenger pathways in post-synaptic neurons and inhibits G-proteins and cyclic AMP formation(1, 2). Increased G-protein coupled receptors have been shown in post-mortem studies of patients with BD compared with age-matched controls (35). These studies imply that dopaminergic neurotransmission and its genetic variation underscore its significance in the treatment of BD.

Calcium is a cation that plays multiple important roles in cellular functioning, and maintenance of calcium homeostasis is essential for cellular signalling and integrity efficiency (36, 37). However, dysregulation of intracellular calcium has been documented in patients with BD (38). Lithium blocks calcium uptake into cells by attenuating calcium influx after activating NMDA receptors (39). As a result, lithium helps decrease intracellular calcium levels and intracellular calcium stores by blocking calcium uptake into cells after NMDA receptor activation (40). Lithium has been also observed to lead to the depletion of free inositol which is an intracellular signalling molecule that can trigger the release of intracellular calcium thus reducing intracellular stores (6, 41). Genetic studies also reported that calcium-channel genes such as *CACNAID, CACNA1C*, and CACNB3 are associated with lithium response. The expression profile of these genes is amplified by lithium (5). Cellular studies have also shown that lithium reduces the flow of calcium ions and the percentage of signaling neurons, specifically in lithium-responsive neuronal networks (42, 43). These data suggest the importance of calcium channel signaling pathways and their genetic variation to lithium’s therapeutic effects.

Mitochondria are intracellular organelles responsible for the synthesis of cellular energy via the production of adenosine triphosphate (ATP) through oxidative phosphorylation and other cellular processes (44). Brain imaging studies have revealed mitochondrial damage, including abnormal distribution and morphological appearance in patients with BD (45). Genetic analysis on transgenic mice with mitochondrial DNA polymerase mutations and mitochondrial dysfunction has also demonstrated mitochondrial abnormalities may be related BD-like models (24). Studies examining gene and protein expression have shown reduced levels of enzymes involved in ATP production and storage in BD (46). Lithium treatment modulates electron transport chain and oxidative phosphorylation activity (11, 47). In neuronal culture, cells from lithium responsive BD patients show mitochondrial gene expression changes that are associated with neuronal hyperexcitability that is rescued by lithium treatment (8, 48). Lithium has also been reported to increase the life span of mitochondria and correct low mitochondrial oxygen consumption rate (9). These studies exemplify that genetic variation of the mitochondria associates pathways be associated with lithium response in BD.

Circadian rhythm is an important mediator of lithium’s effect on mood related behaviors (49). Disturbance in circadian rhythm is common among BD patients, and lithium was found to enhance the resynchronization of sleep rhythm through modulation of clock gene expression (49, 50). Chronic lithium also modulates the expression of certain clock genes (51) and variants are associated with lithium response (52). Moreover, lithium stimulates the transcription of clock genes through the stimulation of expression promoters (53). These studies imply that the genetic variation in circadian rhythm and GSK pathways could help to predict lithium treatment response in patients with BD.

Lithium has suite effects on several neuroprotective pathways through its direct inhibition of Glycogen synthase kinase 3 (GSK3) (54). GSK3 genes are involved in gene transcription and has an effect on neuroprotective cellular mechanisms that is critical to clinical response to lithium (54). Extracellular signals transduced through lead to phosphorylation and inactivation GSK-3. However, GSK-3 opposes its own phosphorylation through (55): 1) GSK-3 inhibits Akt by binding to and stabilizing a β-arrestin complex that brings Akt into contact with its deactivation phosphatase PP2A. Inactivation of Akt thereby prevents phosphorylation and inhibition of GSK-3. Lithium disrupts this complex (56, 57) allowing Akt to remain active and phosphatase GSK-3. 2) GSK-3 promotes its own dephosphorylation by activating protein phosphatase 1(PP1) which is inhibited by 1-2; GSK-3 phosphorylation of I-2 causes it to dissociate from PP1, which then dephosphorylates GSK-3, restoring GSK-3 activity. Lithium prevents PPI from dephosphorylating GSK-3 in an I-2 dependent manner (58). A study investigating how lithium modulates GSK3β inhibition, revealed that GSK3β gene expression predicted lithium clinical response for manic and depressive symptoms (59, 60). These studies suggest that GSK3β is a well-supported direct target of lithium action in BD, and related pathways are worthy of inclusion in genomics studies of response.

### **Sample selection procedure for the replication cohorts**

PsyCourse is an ongoing multisite, naturalistic longitudinal cohort study involving adults aged 18 years and above, with an ICD-10 lifetime diagnosis of BD I or II, recurrent major depression, schizoaffective disorder, or brief psychotic disorder. Consented participants are included based on clinical assessments and disease registries at participating clinical centers, with diagnosis confirmed using DSM IV criteria (61-63). Individuals were excluded if they did not meet DSM-IV diagnostic criteria, had insufficient language ability or were intellectually impaired. In contrast, the Bipolife cohort was recruited from a German-based nationwide network of university hospitals aiming to improve early recognition and intervention of patients with BD (64). Patients aged between 18 and 35 years with a diagnosis of BD and at least one depressive, manic, or mixed episode during the preceding two years were eligible (64).

### **Genotyping, quality control, and imputation procedures for the replication cohorts**

The genotyping, QC and imputation for the PsyCourse cohort were previously performed as described elsewhere (65, 66). Briefly, genomic DNA was extracted from blood samples and underwent genome-wide genotyping using the Illumina Infinium CoreExome-24+Human PsyChip Consortium version 1,0 and 1.1 (Illumina, USA), yielding approximately 590,000 SNPs (63). Following standard QC procedures and population substructure analysis in PLINK v 1.9 (67), genotype data were imputed to the 1000 Genomics phase 3 reference panel(68) using SHAPEIT2 (69) and IMPUTE2 (70). Post-imputation QC included the removal of SNPs with poor imputation quality (INFO <0.8), a Hardy-Weinberg equilibrium test *p* < 1x10^-6^ and a MAF < 1%. Individuals with a mismatch between information on phenotypic and genotypic sex, genotyping call rate <98%, genetic duplicates, or intra-sample relatedness were excluded from the downstream analyses (63, 65, 66). The genotyping and imputation procedures for the Bipolife cohort were similar to the ConLi^+^Gen cohort (64, 71).

### **Procedures for developing pathway-specific polygenic scores**

For the ConLi^+^Gen cohort, which includes participants from 13 countries, we implemented a widely accepted leave-one-country-out (LOC) procedure (72, 73) where PGSs were calculated for participants of one country at a time (target sample) using GWAS summary statistics from the remaining 12 countries (discovery sample). For instance, to calculate a PGS for Australian participants (target sample), GWAS summary data from all other countries, excluding Australia, was used as the discovery cohort. The advantage of the LOC approach over other methods, such as sample splitting methods, is that it accounts for potential genetic variation based on participants' country of origin, thereby considering geographical differences and ethnic diversity.

Using this LOC approach, we generated 26 sets of GWAS summary statistics (discovery datasets) for both categorical and continuous lithium treatment outcomes. Each GWAS was performed using PLINK version 2.0 adjusting for regression models adjusted for age, sex, chip type, and the first four principal components (PCs). The number of PCs was determined by examining the scree plot of eigenvalues in the principal component analysis, which indicated that the first four PCs can capture the underlying population structure (Supplementary Figure 1)

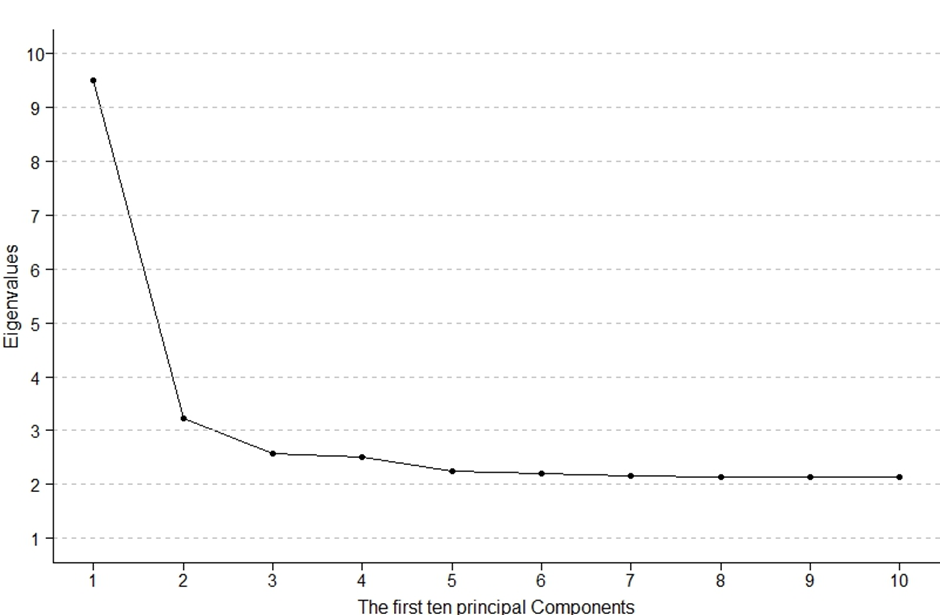

**Supplementary Figure 1:** Scree plot for determining the number of principal components considered in our analysis

Each of the PS_PGSs_ was computed using PGScs method that has shown superior performance over standard methods (74). PGS-cs employs a Bayesian regression framework, incorporating continuous shrinkage priors on effect sizes and accounting for linkage disequilibrium (LD) among SNPs (75). Continuous shrinkage applies a proportionate amount of reduction on the coefficient estimates of SNPs based on their strength of association. This process yields a pruned set of SNPs with their posterior effect sizes for downstream analysis (75). To estimate SNP posterior effect sizes, genetic data from the UK Biobank European subsample was used as an external panel of reference for LD pattern and global shrinkage parameter *(--phi=1e-2*) with other default options. Polygenic scores for each pathway were then computed by summing up the dosages of effect alleles (ranging from 0 to 2) for independent genotyped SNPs (j to k) annotated within a specific pathway, multiplied by their respective posterior SNP weight inferred from PGScs (β) (formula 1).

$$PGSi=\sum_{j}^{k} \beta SNPj x dosage of SNPj$$

………………………………………Formula-1

The above process was conducted for each of the nine pathways and for both continuous and categorical lithium treatment responses. Prior to conducting association analyses, the scores for each pathway underwent standardization to a mean of zero and a standard deviation of one.

### **Duplicate genes across pathways**

**Supplementary Table 3:** Duplicated genes across selected pathways

| Pathways | ACH | CA | CIR | DOP | GABA | GLUT | GSK | MITO | NMDA |
| --- | --- | --- | --- | --- | --- | --- | --- | --- | --- |
| ACH | **164** | 5 | 9 | 51 | 5 | 35 | 2 | 0 | 6 |
| CA | 5 | **134** | 8 | 5 | 8 | 9 | 0 | 9 | 0 |
| CIR | 9 | 8 | **129** | 12 | 7 | 13 | 1 | 4 | 0 |
| DOP | 51 | 5 | 12 | **155** | 7 | 15 | 12 | 4 | 0 |
| GABA | 5 | 8 | 7 | 7 | **76** | 24 | 0 | 0 | 0 |
| GLUT | 35 | 9 | 13 | 15 | 24 | **92** | 1 | 0 | 7 |
| GSK | 2 | 0 | 1 | 12 | 0 | 1 | **18** | 2 | 0 |
| MITO | 0 | 9 | 4 | 4 | 0 | 0 | 2 | **163** | 0 |
| NMDA | 6 | 0 | 0 | 0 | 0 | 7 | 0 | 0 | **11** |

*Abbreviations:* ACH: Acetylcholine; CA: Calcium channel signalling; CIR: Circadian rhythm; DOP: Dopamine; GABA: Gamma-aminobutyric acid; GSK: Glycogen synthase kinase; MITO: Mitochondria; NMDA: N-methyl-D-aspartate

For example, five genes were extracted in calcium channel signaling and acetylcholine pathways. Hence, some genes were duplicated across the nine potential pathways, and we performed a multivariate regression modelling considering only PS_PGSs_ that showed a significant association with lithium response in the univariate model.

### **Comparative analysis with genome-wide lithium responsiveness PGS**

To compare the model performance of PS_PGS_, we developed genome-wide PGS using the LOC procedure employing the genome-wide variants of lithium responsiveness in similar cohorts (4). The polygenic score of lithium responsiveness (Li^+^_PGS_) was calculated for participants of one country at a time (target sample) using GWAS summary statistics from the remaining 12 countries (discovery sample). Association analysis was then performed with lithium treatment response in both continuous and categorical outcomes, adjusted for age, sex, chip type, and the first four PCs. Finally, we compared the combined PS_PGS_ model performance with the genome-wide lithium responsiveness PGS using *r2redux* R package (76). This package calculates test statistics for R^2^ values from PGS models, including their variance and covariance of R^2^ values, p-values, and 95% confidence interval) and tests whether R^2^ values are significantly different regardless of whether the two PGS sets are independent or dependent (76).

We found a significant association between genome-wide Li^+^_PGS_ and lithium treatment response in both categorical (p = 4.13x10^-9^; R^2^ = 2.87%) and continuous (p = 2.41x10^-7^; R^2^ = 2.69%) outcomes using the LOC approach. When comparing the R^2^ of Li^+^_PGS_ and combined PS_PGS_, there was no statistically significant difference for either categorical (R^2^ difference = 84%; p = 0.58, non-nested model) or continuous (R^2^ difference = 0.49%; p = 0.88, non-nested model) lithium treatment responses (Supplementary Table 4).

**Supplementary Table 4:** The results of genome-wide variants of lithium responsiveness PGS and combined pathway-specific PGS using leave-one-country-out analysis.

| PGS | Categorical outcome | | | Continuous ALDA score | | |
| --- | --- | --- | --- | --- | --- | --- |
|  | aOR (95%CI) | *p* | R^2^(%) | β(95%CI) | *p* | R^2^(%) |
| Genome_wide PGS | 1.14(1.03, 1.27) | 4.13x10^-9^ | 2.87 | 0.45(0.31, 0.57) | 2.41x10^-7^ | 2.69 |
| Combined PS_PGS_ | NA | <0.01 | 3.71 | NA | <0.01 | 3.18 |

*Legends:* The analysis models were adjusted for age, sex, chip type, and the first four principal components (PCs). The observed R^2^ for the categorical outcome is transformed to a liability scale. *Combined PS_PGS_* represents a multivariate analysis of pathway-specific PGS with p-values less than 0.05 in the univariate analysis at least either of lithium response i.e., acetylcholine, GABA, calcium signaling, mitochondria, circadian rhythm, and GSK potential pathways. *Genome-wide PGS* represents PGS for genome-wide variants of lithium responsiveness for lithium treatment response in a one-country-leave-out procedure.

*Abbreviations:* aOR=Adjusted odds ratio; R^2^ = variance explained by polygenic scores; *p* = p-value; ALDA = Retrospective Criteria of Long-Term Treatment Response in Research Subjects with Bipolar Disorder; NA = Not applicable

### **The association of pathway-specific polygenic scores and good lithium treatment response a****mong bipolar patients treated with lithium in decile analysis**

**Supplementary Table 5:** The association of pathway-specific polygenic scores and good lithium treatment response among bipolar patients treated with lithium in decile analysis

| PGS deciles | Acetylcholine | | GABA receptor | | Mitochondria | | Calcium channel | GSK | Circadian rhythm |
| --- | --- | --- | --- | --- | --- | --- | --- | --- | --- |
|  | Categorical outcome | Continuous outcome (β) | Categorical outcome | Continuous outcome | Categorical outcome | Continuous outcome | Continuous outcome | Continuous outcome | Continuous outcome |
| 1^st^ | 1.00 | 0.00 | 1.00 | 0.00 | 1.00 | 0.00 | 0.00 | 0.00 | 0.00 |
| 2^nd^ | 1.18(0.73,1.90) | 0.45(-0.09,0.99) | 1.04(0.67,1.65) | 0.76(0.21,1.30)* | 0.75(0.49,1.12) | -0.37(-0.92,0.17) | 0.17(-0.38,0.71) | 0.62(0.08,1.16) | 0.12(-0.42,0.66) |
| 3^rd^ | 1.27(0.79,2.03) | 0.71(0.16,1.26)* | 1.25(0.80,1.95) | 0.78(0.24,1.33)* | 0.81(0.54,1.22) | -0.39(-0.93,0.16) | 0.73(-0.38,1.28) | 0.15(-0.40,0.70) | 0.01(-0.53,0.55) |
| 4^th^ | 1.01(0.63,1.63) | 0.59(0.05,1.14)* | 1.56(1.01,2.41)* | 0.62(0.07,1.16)* | 0.71(0.47,1.08) | -0.32(-0.87,0.23) | 0.47(0.18,1.02)* | 0.80(0.26,1.35)* | -0.06(-0.60,0.45) |
| 5^th^ | 1.32(0.83,2.10) | 0.78(0.23,1.32)* | 1.41(0.91,2.19) | 0.62(0.07,1.16)* | 0.63(0.41,0.95)* | -0.50(-1.04,0.05) | 0.50(-0.05,1.05) | 0.53(-0.02,1.08) | 0.38(-0.16,0.93) |
| 6^th^ | 1.37(0.85,2.20) | 1.09(0.54,1.64)* | 1.42(0.88,2.05) | 0.49(-0.06,1.04) | 0.66(0.43,0.99)* | -0.73(-1.28,-0.18)* | 0.66(0.12,1.21)* | 0.33(-0.22,0.88) | 0.11(-0.44,0.65) |
| 7^th^ | 1.81(1.15,2.86)* | 1.13(0.58,1.67)* | 1.53(0.98,2.38) | 0.25(-0.30,0.80) | 0.56(0.36,0.85)* | -0.59(-1.13,-0.04)* | 0.30(-0.25,0.85) | 0.49(-0.06,1.03) | 0.31(-0.24,0.85) |
| 8^th^ | 2.07(1.31,3.25)* | 1.10(0.55,1.64)* | 1.48(0.95,2.30) | 0.70(0.15,1.25)* | 0.57(0.38,0.88)* | -0.31(-0.86,0.24) | 0.85(0.30,1.40)* | 0.97(0.42,1.52)* | 0.71(0.16,1.25)* |
| 9^th^ | 2.17(1.38,3.41)* | 1.14(0.59,1.69)* | 1.37(0.88,2.14) | 0.77(0.22,1.32)* | 0.55(0.36,0.85)* | -0.54(-1.09,0.04) | 0.67(0.12,1.21)* | 0.97(0.42,1.52)* | 0.58(0.04,1.13)* |
| 10^th^ | 3.03(1.95,4.69)* | 1.62(1.07,2.16)* | 2.01(1.30,3.09)* | 0.77(0.22,1.32)* | 0.52(0.33,0.80)* | -1.03(-1.58,-0.48)* | 0.73(0.18,1.27)* | 0.79(0.24,1.34)* | 0.35(-0.19,0.01) |

*Legends:* A statistically significant association is determined at *p* < 0.05, adjusted for age, sex, chip type and the first four genetic principal components. (*) denotes statistical significant in decile comparison

*Abbreviations*: GABA: Gamma-aminobutyric acid; GSK: Glycogen synthase kinase

### **Sensitivity analysis**

To evaluate the robustness and reliability of our findings, we developed PSPGS for randomly selected biological pathways (melanin and keratinocyte differentiation pathways) which have a limited implication in the pharmacokinetics of lithium action. The association of these PS_PGS_ with lithium response was tested, and the results are discussed accordingly. No significant associations were observed between the PGSs for the melanin and keratinocyte differentiation pathways and lithium treatment response both in categorical or continuous scales in patients with bipolar disorder (p > 0.1). The supplementary table 6 presents the association analyses of PS_PGSs_ for these random pathways with lithium treatment response.

**Supplementary Table 6:** Sensitivity analysis on polygenic score for random pathway and lithium treatment response.

| Pathways | Genes (n) | SNPs (n) | Categorical ALDA outcome | | | Continuous ALDA score | | |
| --- | --- | --- | --- | --- | --- | --- | --- | --- |
|  |  |  | aOR (95%CI) | *p* | R^2^(%) | β(95%CI) | *p* | R^2^(%) |
| Keratinocyte differentiation | 62 | 2459 | 1.08(0.98, 1.20) | 0.26 | 0.21 | 0.13(-0.02, 0.21) | 0.10 | 0.01 |
| Melanin | 73 | 2358 | 1.02(0.93, 1.13 | 0.71 | 0.01 | 0.08(-0.04, 0.21) | 0.53 | 0.01 |

*Legends:* A statistically significant association is determined at p < 0.05, adjusted for age, sex, chip type and the first four genetic principal components.
